# Occupation, Worker Vulnerability, and COVID-19 Vaccination Uptake: Analysis of the Virus Watch prospective cohort study

**DOI:** 10.1101/2022.06.12.22276307

**Authors:** Sarah Beale, Rachel Burns, Isobel Braithwaite, Thomas Byrne, Wing Lam Erica Fong, Ellen Fragaszy, Cyril Geismar, Susan Hoskins, Jana Kovar, Annalan M D Navaratnam, Vincent Nguyen, Parth Patel, Alexei Yavlinsky, Martie Van Tongeren, Robert W Aldridge, Andrew Hayward, the Virus Watch Collaborative

**Affiliations:** Centre for Public Health Data Science, Institute of Health Informatics, University College London, UK, NW1 2DA; Institute of Epidemiology and Health Care, University College London, London, UK, WC1E 7HB; Department of Infectious Disease Epidemiology, London School of Hygiene and Tropical Medicine, Keppel Street, London, UK, WC1E 7HT; Centre for Occupational and Environmental Health, University of Manchester, Manchester, UK, M13 9PL

## Abstract

**Background:** Occupational disparities in COVID-19 vaccine uptake can impact the effectiveness of vaccination programmes and introduce particular risk for vulnerable workers and those with high workplace exposure. This study aimed to investigate COVID-19 vaccine uptake by occupation, including for vulnerable groups and by occupational exposure status.

**Methods:** We used data from employed or self-employed adults who provided occupational information as part of the Virus Watch prospective cohort study (*n*=19,595) and linked this to study-obtained information about vulnerability-relevant characteristics (age, medical conditions, obesity status) and work-related COVID-19 exposure based on the Job Exposure Matrix. Participant vaccination status for the first, second, and third dose of any COVID-19 vaccine was obtained based on linkage to national records and study records. We calculated proportions and Sison-Glaz multinomial 95% confidence intervals for vaccine uptake by occupation overall, by vulnerability-relevant characteristics, and by job exposure.

**Findings:** Vaccination uptake across occupations ranged from 89-96% for the first dose, 87-94% for the second dose, and 75-86% for the third dose, with transport, trade, service and sales workers persistently demonstrating the lowest uptake. Vulnerable workers tended to demonstrate fewer between-occupational differences in uptake than non-vulnerable workers, although clinically vulnerable transport workers (76%-89% across doses) had lower uptake than several other occupational groups (maximum across doses 86-96%). Workers with low SARS-CoV-2 exposure risk had higher vaccine uptake (86%-96% across doses) than those with elevated or high risk (81-94% across doses).

**Interpretation:** Differential vaccination uptake by occupation, particularly amongst vulnerable and highly-exposed workers, is likely to worsen occupational and related socioeconomic inequalities in infection outcomes. Further investigation into occupational and non-occupational factors influencing differential uptake is required to inform relevant interventions for future COVID-19 booster rollouts and similar vaccination programmes.

## Introduction

COVID-19 vaccination has become a cornerstone of the global pandemic response since the licensing of safe and effective vaccines in December 2020. Licensed COVID-19 vaccines minimise symptomatic SARS-CoV-2 infection risk and greatly reduce the risk of severe morbidity and mortality from COVID-19 (1,2). However, the efficacy of vaccination programmes at reducing individual and population-level risk from COVID-19 depends on high levels of uptake across the eligible population. Understanding disparities in vaccination uptake across population subgroups is consequently essential to inform vaccination strategies in the context of the current phase of the pandemic and to provide background information for future COVID-19 vaccination drives and other vaccine-modifiable outbreak scenarios.

In working-age adults, the workplace is a persistent and notable source of potential SARS-CoV-2 exposure (3–6), with considerable variation across occupational groups. Occupations that cannot be completed from home and involve high levels of contact with others - such as healthcare, teaching, public transport and personal service occupations - are associated with elevated infection risk in studies across a range of global regions (3,7–14). Differential vaccine uptake in high-exposure occupations is an area of concern, particularly for workers with conditions that render them vulnerable to severe COVID-19. As well as being an important source of potential exposure, occupation may influence vaccine uptake itself. This influence may be direct, by creating structural barriers or enablers for vaccination, e.g. time off work to receive vaccination or promotion programmes in the workplace, and via workplace cultures. It may also operate indirectly through the influence of non-work factors related to occupation that may influence vaccination-related attitudes and behaviours, such as socio-economic position. Investigating vaccine uptake by occupation is consequently important due to its influence on exposure and potential influence on vaccination-related behaviour.

Preliminary evidence from the United States (USA) and United Kingdom (UK) suggests differential COVID-19 vaccine uptake by occupation. An early USA-based study found higher first dose vaccination uptake in the initial phase of COVID-19 vaccine availability between January-March 2021 amongst healthcare, science, education, and managerial occupations (>40% vaccinated), and lower vaccine uptake in manual occupations including construction and extraction workers (18% vaccinated) (15). Similar trends were identified via linkage between UK national vaccination records and 2011 occupational census data for later doses. Completion of two-dose vaccination courses for workers between 40-64 years of age was highest for managerial and professional occupations (>90%), and lowest for elementary occupations – including manufacturing, processing, and construction workers and waiters (all <80%) (16). Occupations classified as least able to work remotely tended to demonstrate lower vaccine uptake than occupations most able to work remotely (16). Similar findings were identified regarding uptake of third booster doses up to 28 February 2022 in the UK, which included workers over 18 and linkage to 2021 census data (17). Health and education professionals had the highest levels of three-dose booster uptake (respectively 85% and 84%) and elementary trade, sales and service occupations the lowest levels (58%-65%). These findings indicate marked occupation differences in vaccination uptake across doses, with greater uptake in professional occupations and lower uptake in trade and service occupations. However, trends from single studies with particular sample-related or linkage-related concerns may not generalise across other populations and time periods. Investigation into the relationship between exposure risk and vaccination uptake, as conducted in the UK for the second vaccination dose, is also indicated for other doses.

Additionally, evidence of differential uptake across occupations raises concerns regarding vaccination uptake and consequent protection for vulnerable workers. Notable occupational differences in the prevalence of conditions associated with greater risk of severe COVID-19 - including cardiometabolic and lung diseases and obesity - were well-established pre-pandemic (18)((19)(20). Notably, these conditions tend to be particularly prevalent amongst occupational groups identified in early studies to have lower COVID-19 vaccine uptake, such as public transport and manufacturing occupations. Given that this subpopulation of workers may be particularly impacted by differential uptake, triangulation of workers’ vaccination uptake with factors influencing their vulnerability status is warranted.

### Aims and Objectives

This study aimed to investigate how vaccination status and clinical vulnerability varied across occupations and interacted with one another based on data from Virus Watch, a large prospective cohort study in England and Wales (21). Objectives were:

1. To investigate vaccination uptake for the first, second, and third dose of COVID-19 vaccine by occupation
2. To investigate how vaccination uptake amongst vulnerable workers varied by occupation for each dose of COVID-19 vaccine
3. To investigate how vaccination uptake for each dose varied by potential occupational exposure to SARS-CoV-2

## Methods

### Ethics Approval

The Virus Watch study was approved by the Hampstead NHS Health Research Authority Ethics Committee: 20/HRA/2320, and conformed to the ethical standards set out in the Declaration of Helsinki. All participants provided informed consent for all aspects of the study.

### Participants

Participants in the current study were an adult sub-cohort of the Virus Watch cohort enrolled prior to 12/02/2022 (see Figure 1), the cut-off date for study recruitment. Participants who met the following criteria were included in the present study (1) adult ≥18 years, (2) employed or self-employed full-time or part-time and reported their occupation upon study registration, (3) provided information on vaccination status recorded via linkage or self-report (see Outcomes below).

**Figure 1.**
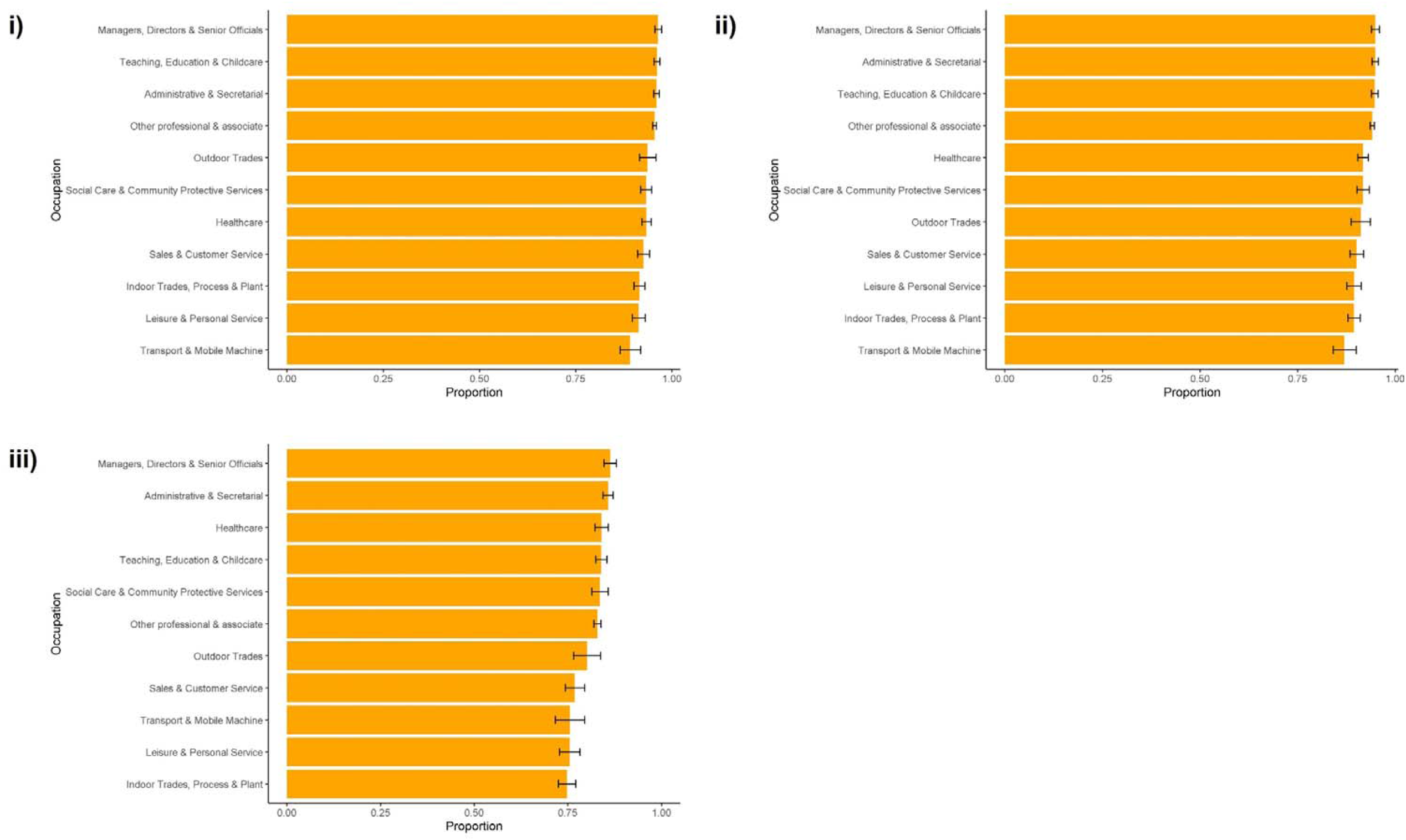
Proportions and 95% Confidence Intervals for Vaccination Status by Occupation for First Dose (i), Second Dose (ii), and Third Dose (iii)

### Exposure

#### Occupation

Participants provided their occupation as free text at study registration, which was coded into UK Standard Occupational Classification (SOC) 2020 codes (22) using semi-automatic processing in Cascot Version 5.6.3 (22,23). Occupational groups were then classified as follows for the present study based on SOC codes to broadly reflect occupational environments while preserving, where possible, ONS-derived occupational groupings (see (24) for further details of occupational classification): administrative and secretarial occupations; healthcare occupations; indoor trade, process & plant occupations; leisure and personal service occupations; managers, directors, and senior officials; outdoor trade occupations; sales and customer service occupations; social care and community protective services; teaching education and childcare occupations; transport and mobile machine operatives; and other professional and associate occupations (broadly office-based, non-essential professional occupations). See Supplementary Table 1 for SOC codes within each category.

**Table 1.** Demographic Features of Study Participants

| Characteristic | n (%) (N = 19,595) |
| --- | --- |
| Age |  |
| <30 | 1,723 (8.8%) |
| 30-39 | 3,378 (17%) |
| 40-49 | 4,288 (22%) |
| 50-59 | 5,512 (28%) |
| 60+ | 4,693 (24%) |
| Unknown | 1 |
| Sex |  |
| Female | 10,902 (56%) |
| Male | 8,658 (44%) |
| Missing | 34 (0.2%) |
| Unknown | 1 |
| Vaccination Status |  |
| Unvaccinated | 1,070 (5.5%) |
| One Dose | 300 (1.5%) |
| Two Doses | 2,102 (11%) |
| Three Doses | 16,123 (82%) |
| Clinically Vulnerable | 6,801 (35%) |
| Obese | 3,983 (24%) |
| Unknown | 2,919 |
| Ethnicity |  |
| White British | 15,863 (82%) |
| White Other | 1,821 (9.4%) |
| Mixed | 336 (1.7%) |
| South Asian | 892 (4.6%) |
| Other Asian | 196 (1.0%) |
| Black | 209 (1.1%) |
| Other Ethnicity | 113 (0.6%) |
| Unknown | 165 |
| Region |  |
| East Midlands | 1,739 (8.9%) |
| East of England | 3,790 (19%) |
| London | 3,491 (18%) |
| North East | 864 (4.4%) |
| North West | 1,963 (10%) |
| South East | 3,733 (19%) |
| South West | 1,321 (6.7%) |
| Wales | 487 (2.5%) |
| West Midlands | 1,012 (5.2%) |
| Yorkshire and The Humber | 930 (4.7%) |
| Unknown | 265 |

#### Work-Related Exposure Risk

Work-related exposure risk was derived based on the Job Exposure Matrix (25), a six-dimension measure classifying the occupational risk of SARS-CoV-2 based on SOC-2020 scores. Occupations are allocated a score between 0 (no risk) and 3 (high risk); a mean JEM score was calculated for each occupation rounded up to the nearest integer, with a mean score between <=1 denoting low risk, 2 denoting elevated risk, and 3 denoting high risk in the current analyses. Work-related exposure risk was not further stratified by occupation as several occupational groups contained no workers in either the low risk and/or high risk groups.

### Outcome

#### Vaccination Status

The primary outcome of interest was binary vaccination status for each of the three doses of COVID-19 vaccine recommended for all adults in the UK (yes/no received first, second, and third dose) (24,26). While some clinically extremely vulnerable adults have been recommended to receive a fourth dose of a COVID-19 vaccine, only the first three doses were considered in this analysis due to their applicability to the entire working age population. Data were cut on 21/04/2022, with vaccine doses up to this date recorded; this date occurred several months after appointments for third (‘booster’) doses of COVID-19 vaccine were offered to all adults over 18 in the UK (31/12/2021).

Vaccination status was derived from the following sources in order of preference: (1) linkage to the UK National Immunisation Management Service (NIMS), which contains officially-reported records of NHS COVID-19 vaccinations (available for vaccinations between 9 October 2020 – 23 December 2021), or (2) self-reported vaccinations obtained from responses to Virus Watch weekly surveys (available for full study period). Self-reported vaccinations were asked retrospectively in the Virus Watch weekly survey from 11 – 18 January 2020 to record any vaccinations prior to this date, and subsequently were asked weekly from 25 January 2020 onwards. If participants had a second or third dose recorded but were missing information about earlier dose(s) (*n*=1470 for dose 1 and *n*=599 for dose 2) then they were considered to have received previous doses.

### Stratification Variables

#### Clinical Vulnerability

Clinical vulnerability status was binary coded (not vulnerable versus vulnerable/extremely vulnerable) based on classifications set out by Public Health England/UK Health Security Agency, the Department of Health and Social Care, and the Joint Committee on Vaccination and Immunisation (27,28) to denote vulnerability to severe COVID-19 morbidity and mortality based on the presence of specific medical conditions. Participants indicated whether they suffered from a range of medical conditions deemed by UK health authorities to denote clinical vulnerability at registration and in monthly surveys collected in February and May 2021 to record any new-onset conditions in long-term participants.

Given that vulnerability was investigated in the context of current vaccination status, it was derived based on the presence of any qualifying condition recorded at any point. Conditions denoting vulnerability or extreme clinical vulnerability were collapsed into a single category due to sample size limitations. Please see (29) for further details of vulnerability classification within Virus Watch.

#### Obesity

Obesity was included as a binary variable (obese versus not obese) based on body mass index (BMI>=30) following UK NHS classification (30). Obesity is associated with higher risk of severe COVID-19 morbidity and mortality (31,32). Participants provided their height and weight upon study registration, which was used to calculate body mass index.

#### Age

Age was included as a binary variable, with younger (<60) and older (>60) age groups to reflect increased risk of severe COVID-19 outcomes in older people (31,32) while conserving statistical power.

### Statistical Analyses

To investigate patterns of differential vaccination status and vulnerability by occupation and facilitate comparison between all occupational groups, proportions and Sison-Glaz confidence intervals for multinomial proportions (33) were presented by occupation for: vaccination uptake for each dose, clinical vulnerability status, and vaccination uptake for each dose stratified by clinical vulnerability, obesity, age, and exposure risk. Vaccination uptake on time for each dose was not further stratified by vulnerability-relevant characteristics due to smaller cell sizes than vaccination uptake overall.

Given the crucial role of vaccination in reducing likelihood of severe COVID-19 outcomes, this analysis was concerned with identifying patterns of differential vaccine uptake and vulnerability to identify at-risk occupations rather than isolating the direct effect of occupation on vaccination status. Consequently, estimates were stratified by vulnerability-relevant factors rather than adjusted. Presentation of proportions and confidence intervals facilitates direct comparison across occupational groups rather than comparison to a reference category.

Missing data for exposure and stratification variables were sparse (<1%) for all variables except obesity (15%) (see Table 1) and complete case analysis was performed for each analysis due to the descriptive approach.

## Results

The present study comprised 19,595 participants, with demographic features reported in Table 1 and participant selection in Supplementary Figure 1.

### Vaccination Status by Occupation

Differences in vaccination uptake emerged by occupation (Figure 1). For the first dose, uptake ranged between 89.1% (95% CI: 86.5%, 91.9%) in transport workers to 96.3% (95.5%, 97.2%) in managerial occupations. Confidence intervals indicated that the four groups with the highest uptake - managerial, administrative, teaching, and other professional occupations - had greater uptake than all other occupations. Patterns were similar for second dose uptake, which ranged from 86.9% (84.1%, 90.0%) in transport workers to 94.9% (93.9%, 95.9%) in managerial occupations. The lowest four groups - transport, indoor trades and process/plant, leisure and personal service, and sales and customer service workers - had lower uptake than all other occupations except outdoor trades. These groups also had lower third dose uptake, which ranged from 74.8% (72.5%, 77.2%) in indoor trades to 86.3% (84.7%, 88.0%) in managerial occupations.

### Vaccination Status and Worker Vulnerability

#### Vulnerability-Related Factors by Occupation

Health-related factors relevant to vulnerability also differed by occupation. The proportion of workers with a condition rendering them clinically vulnerable to severe COVID-19 (Supplementary Figure 2a) ranged from 29.6% (27.2-32.2%) amongst indoor trade, process and plant occupations to 38.8% (35.8-42.0%) in sales and customer service occupations. The proportion of workers living with obesity (Supplementary Figure 2b) ranged from 20.2% (19.2-21.3%) in other professional and associate occupations to 31.7% (27.0-36.8%) in transport and mobile machine operatives. The proportion of older workers over 60 years of age (Supplementary Figure 2c) ranged from 19.6% (18.7%, 20.6%) in other professional occupations to 43.8% (39.2%, 48.6%) in outdoor trade occupations. Confidence intervals for the occupations with the lowest levels of vulnerability-relevant factors did not overlap with those with the highest estimates.

**Figure 2.**
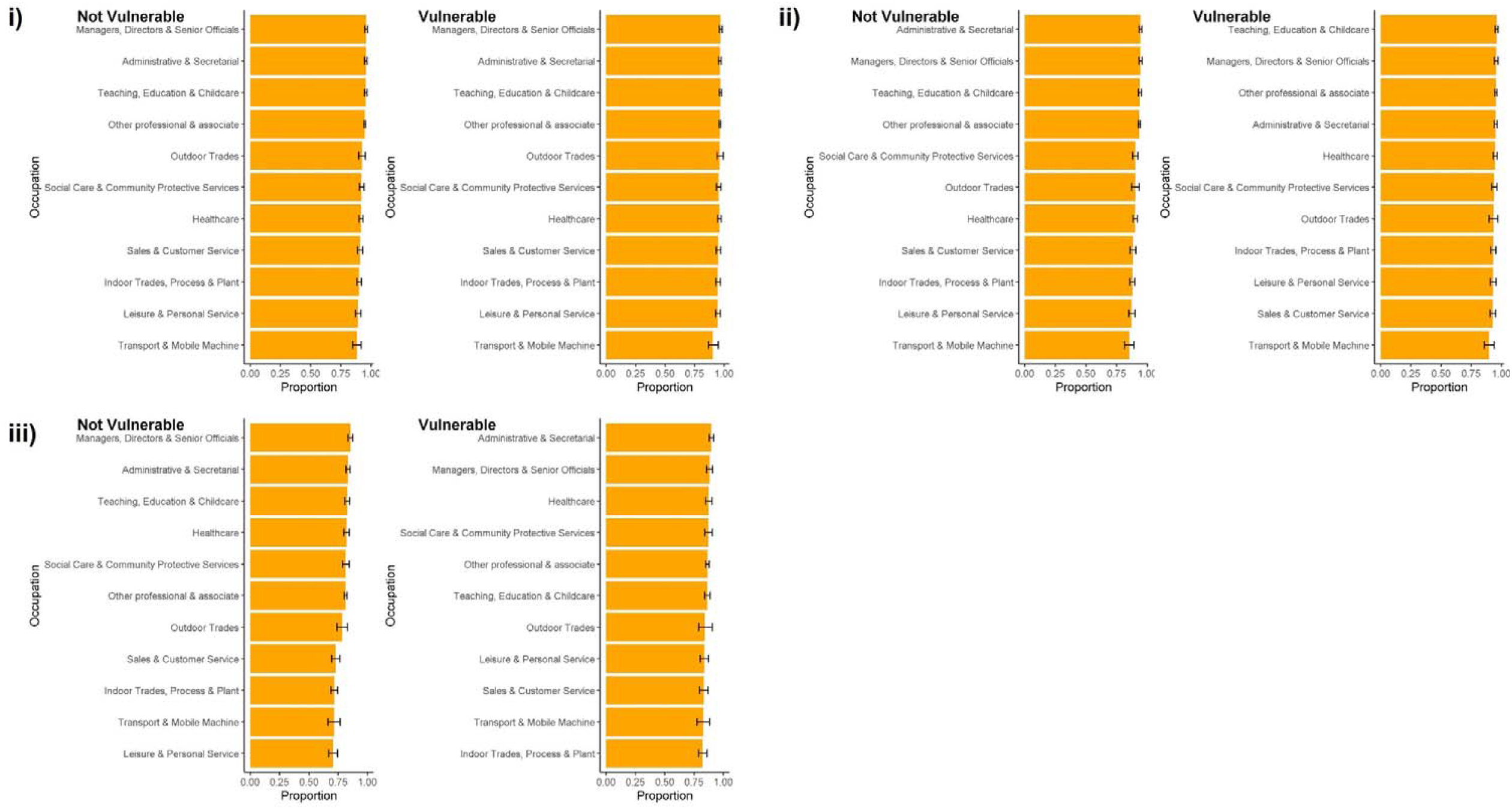
Proportions and 95% Confidence Intervals for Vaccination Status by Occupation, Stratified by Clinical Vulnerability for First Dose (i), Second Dose (ii), and Third Dose (iii)

#### Workers’ Vaccination Status by Vulnerability-Related Factors

Among clinically vulnerable workers, >90% of participants received their first and second dose of COVID-19 vaccine across occupations and >80% received their third dose (Figure 2). Confidence intervals overlapped for all groups except transport occupations, who had consistently lower estimates compared to the groups with the highest uptake for each dose. Between-occupational differences for non-vulnerable participants followed similar patterns to those reported for the full cohort. Similar results were obtained when vaccination uptake was stratified by obesity status (Supplementary Figure 3), with no substantial between-occupational differences identified for workers with obesity and similar patterns to the overall cohort for non-obese participants.

**Figure 3.**
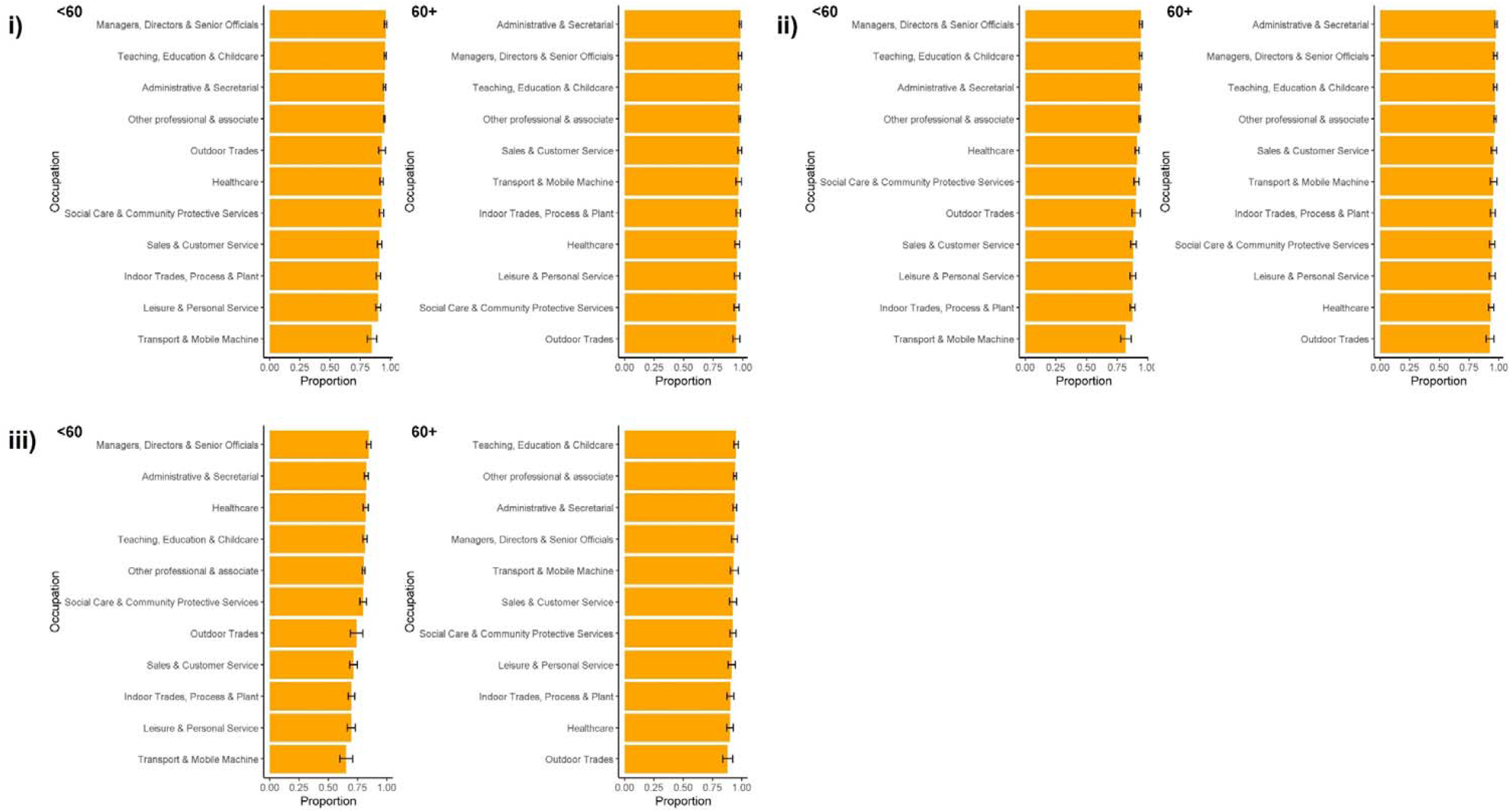
Proportions and 95% Confidence Intervals for Vaccination Status by Occupation, Stratified by Age for First Dose (i), Second Dose (ii), and Third Dose (iii)

Older workers demonstrated high vaccination uptake for the first (>94%), second (>92%), and third (>88%) doses (Figure 3). Confidence intervals overlapped substantially across groups for the first dose. For the second and third doses, the groups with the lowest uptake (outdoor trades and healthcare for both doses, and additionally indoor trades for the third dose) had evidence of lower uptake compared to the occupational groups with the highest estimates. As with clinical vulnerability, between-occupational differences in vaccination uptake was more pronounced for younger workers and were similar to those reported for the whole cohort.

### Workers’ Vaccination Status by Occupations SARS-CoV-2 Exposure

Across all three doses, workers in low-risk occupations demonstrated higher vaccination uptake than those in elevated-risk or high-risk occupations (Figure 4) based on confidence intervals for the proportions. Confidence intervals overlapped between the elevated risk and high-risk groups for all doses.

**Figure 4.**
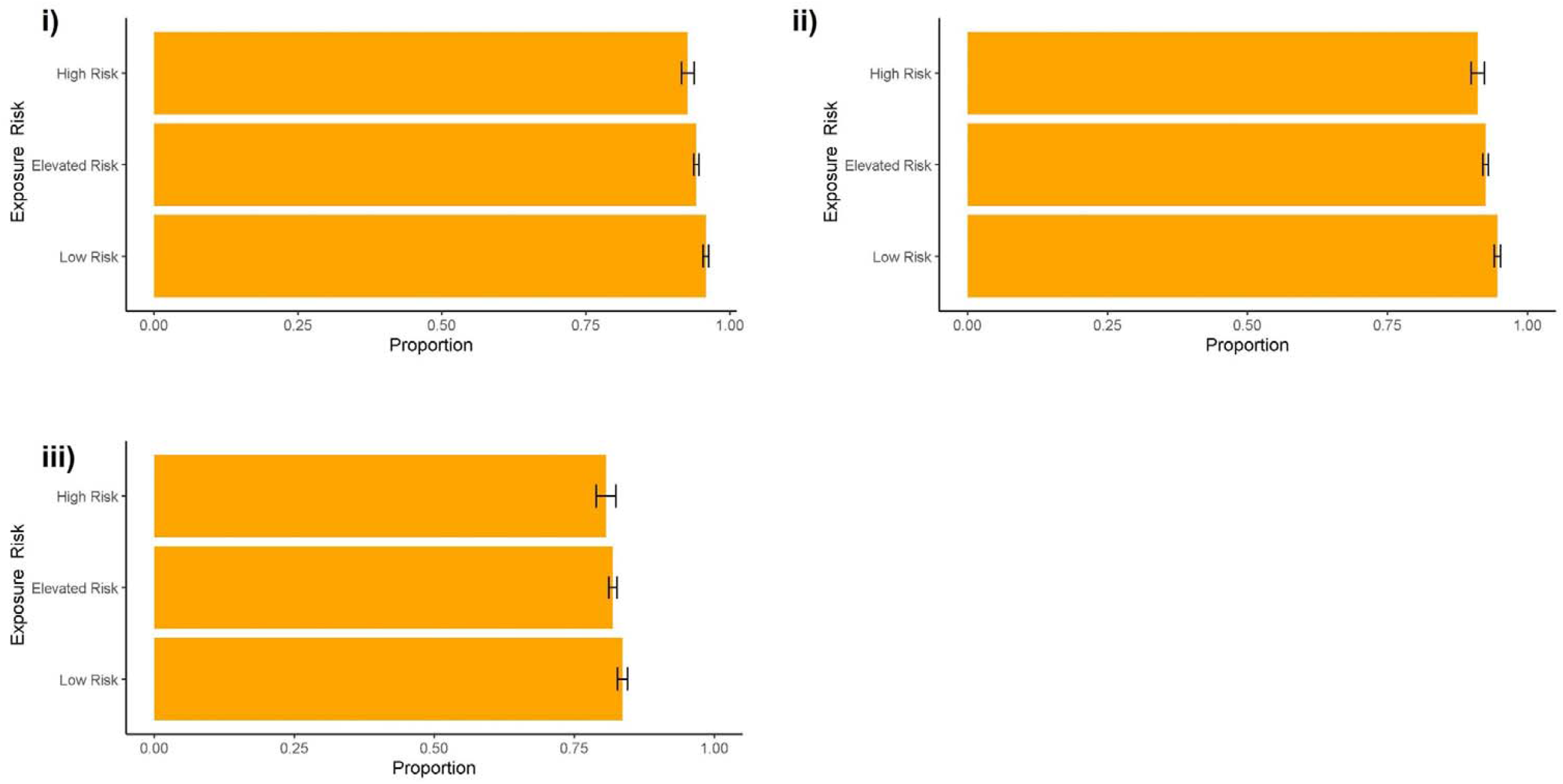
Proportions and 95% Confidence Intervals for Vaccination Status by Job Exposure Risk for First Dose (i), Second Dose (ii), and Third Dose (iii)

## Discussion

Based on a large cohort of workers in England and Wales, we found high (>70%) COVID-19 vaccination uptake across all three recommended doses with variability between occupations, both overall and according to vulnerability-relevant characteristics. Uptake tended to be highest in professional, administrative, and managerial occupations and lowest in trade, transport, leisure and service, and sales occupations. These findings corroborate earlier studies in the USA and UK which found similar gradients in uptake between manual and service occupations and professional occupations (15–17). These between-occupational patterns broadly correspond with occupation-based classification of socio-economic status in the UK (34) and likely reflect psychosocial and structural factors both directly and indirectly related to occupation that influence vaccine-related attitudes and behaviours. Understanding and addressing these factors within affected population groups - including occupation-related factors such as workplace vaccination attitudes and access to paid time off to attend appointments - is an important area for further investigation to reduce vaccination-related inequalities and consequent inequalities in illness outcomes.

Occupational groups that tended to have lower vaccination uptake also tended to demonstrate relatively high levels of workers with vulnerability-relevant factors, such as conditions leading to clinical vulnerability to severe COVID-19 (for sales and leisure and personal service occupations), obesity (for transport and sales occupations), and higher proportions of older workers (for outdoor trade and transport occupations). When vaccination uptake was stratified by vulnerability-relevant factors, however, fewer between-occupational differences emerged in uptake for vulnerable workers than for non-vulnerable ones. This finding may reflect high motivation to receive vaccination due to perceived risk amongst vulnerable workers, as well as targeting of vulnerable groups by the UK national vaccination programme. While high and relatively uniform uptake amongst vulnerable workers is encouraging, exposure to a high proportion of unvaccinated coworkers may introduce increased risk to vulnerable workers in these workplaces due to the role of vaccines in reducing infection and possibly effective transmission (35–37) and poorer vaccine response amongst some vulnerable groups (38,39).

Additionally, some notable occupational differences in uptake emerged for vulnerable participants. In particular, clinically vulnerable transport workers had lower uptake for all doses and older healthcare and trade workers had lower uptake for the second and third doses. Transport workers - identified across previous literature as being at greater risk of SARS-CoV-2 infection and COVID-19 mortality (3,7–14) - present a particular group of interest for future investigation, given that they demonstrated lower overall uptake, higher levels of several vulnerability-relevant factors (obesity and older age), and lower uptake amongst clinically vulnerable participants across doses. Both frontline healthcare workers and older people were prioritised for COVID-19 vaccination in the UK national programme, so lag in uptake amongst this subgroup is also an area of concern, particularly given that later doses are important to render full protection. Nurses and nursing assistants, who have demonstrated greater hesitancy and lower vaccine uptake than medical practitioners in some prior studies (40–42), comprised two of the largest occupational subgroups within the healthcare worker category and may have influenced this finding. Given the high exposure risk in these groups and their proximity to potentially vulnerable patients, further investigation into the reasons underlying lower uptake in this group is recommended in order to understand and improve uptake amongst this targeted population for future vaccination drives.

For all three doses, workers in occupations that were scored as low-risk for SARS-CoV-2 exposure had higher vaccine uptake than those in positions scored as elevated or high risk. These findings correspond to those from the UK Office for National Statistics indicating that workers with a greater ability to work from home demonstrated higher second-dose vaccine uptake (16). Lower vaccination uptake in high-exposure occupations may have impacted the effectiveness of work-from-home guidance for reducing severe morbidity and mortality once vaccines were widely available (16). To reduce inequalities in vaccination uptake and consequent outcomes, future investigation to understand and address occupational and non-occupational factors driving lower uptake in high-risk occupations is warranted.

### Strengths and Limitations

Strengths of the study include the large, occupationally-diverse cohort with linkage to national vaccination records that covered the full three-dose vaccination schedule recommended for adults in the UK. Due to the collection of occupational and health-related information as part of the Virus Watch study, we were able to triangulate occupation, vulnerability, and exposure with vaccination status.

Key limitations of this study include that the Virus Watch cohort is not representative of the working population of England and Wales, with an older, more highly-vaccinated study population with a relatively high proportion of vulnerable workers. The cohort comprised higher numbers of participants in professional and administrative occupations relative to trade, transport, and leisure and service occupations. Self-selection into the cohort likely biased estimates of vaccination uptake upwards across occupational groups, though the effect on between-occupational differences is unclear. Estimates of uptake were higher than those for England and Wales overall, but followed similar trends in terms of higher uptake for the first and second relative to the third dose (43). Some occupational groups had low sample sizes, particularly for vulnerable subgroups, which may have impacted the precision of estimates. Due to sample size limitations, we could not investigate uptake for specific occupations. Similarly, we had to collapse participants classified as clinically vulnerable and clinically extremely vulnerable into a single category, possibly masking differences in uptake depending on the degree of vulnerability. Additionally, linking vaccination uptake by occupation to severe morbidity, mortality, or long-term illness outcomes was beyond the scope of the study and limited by sample size. Missing responses for obesity were likely missing not at random due to weight-related stigma, possibly biassing responses for the obese group.

### Conclusions

This study found evidence of occupational differences in uptake across the first, second, and third dose of COVID-19 vaccine based on a large prospective cohort in England and Wales. Transport, trade, leisure and personal service, and sales occupations tended to have lower vaccine uptake than professional and managerial occupations, in line with previous studies. Occupational differences appeared to be driven primarily by non-vulnerable workers, with fewer differences observed amongst vulnerable workers. However, clinically vulnerable transport workers (of all ages) and healthcare and trade workers over the age of sixty had evidence of lower uptake relative to other groups. Understanding the reasons underlying lower uptake amongst vulnerable workers in these occupational groups is important to inform future COVID-19 booster programmes and other vaccination drives. Workers with low SARS-CoV-2 exposure risk had greater vaccination uptake relative to those with higher risk across all doses, potentially impacting the effectiveness of work-from-home recommendations in the presence of population vaccination programmes and indicating the need to understand determinants of vaccination uptake in groups with high risk of exposure. Transport workers demonstrated lower uptake overall and when stratified by vulnerability, representing an important group for further investigation and intervention to promote vaccine uptake. Vaccination campaigns must consider the barriers facing at-risk groups - including occupational and wider socioeconomic barriers - to ensure equitable coverage amongst population groups with the highest levels of need. Effective interventions to promote uniformly high vaccination uptake amongst vulnerable workers and workers with high levels of work-related exposure will reduce severe morbidity, mortality and long-term illness and provide a blueprint to inform future vaccination drives.

## Data Availability

We aim to share aggregate data from this project on our website and via a "Findings so far" section on our website - https://ucl-virus-watch.net/. We also share some individual record level data on the Office of National Statistics Secure Research Service. In sharing the data we will work within the principles set out in the UKRI Guidance on best practice in the management of research data. Access to use of the data whilst research is being conducted will be managed by the Chief Investigators (ACH and RWA) in accordance with the principles set out in the UKRI guidance on best practice in the management of research data. We will put analysis code on publicly available repositories to enable their reuse.

