## Supplementary Materials for "Occupation, Worker Vulnerability, and COVID-19 Vaccination Uptake: Analysis of the Virus Watch prospective cohort study"

**Supplementary Table 1. UK Standard Occupational Classification 2020 (SOC-2020) Codes within Virus Watch Occupational Categories**

| Virus Watch Occupational Category | UK SOC-2020 Codes | Three Most Prevalent Occupations* (SOC-2020 Unit Group) |
|---|---|---|
| Administrative & Secretarial Occupations | 4111-4217, 9211, 9219, 9233 | 1. Other administrative occupations n.e.c. (22%, n= 553)<br>2. Book-keepers, payroll managers, and wage clerks (10%, n=252)<br>3. Office managers (7%, n=175) |
| Healthcare Occupations | 2211-2259, 3211-3219, 3240, 6131-6133 | 1. Other nursing professionals (22%, n=345)<br>2. Generalist medical practitioners (11%, n=167)<br>3. Nursing auxiliaries and assistants (6%, n=96) |
| Indoor Trades, Process & Plant Occupations | 5211-5250, 5315-5317, 5321-5323, 5411-5449, 8111-8149, 8160, 9131-9139, 9241-9259 | 1. Warehouse operatives (10%, n=142)<br>2. Metalworking production and maintenance fitters (8%, n=103)<br>3. Chefs (7%, n=94) |
| Leisure & Personal Service Occupations | 1221-1225, 1252, 1253, 1256, 1257, 6121, 6129, 6211-6250, 9221-9229, 9231, 9261-9269 | 1. Cleaners and domestics (16%, n=152)<br>2. Kitchen and catering assistants (8%, n=77) |

| | | |
|---|---|---|
| | | 3. Hairdressers and barbers (7%, n=70) |
| Managers, Directors & Senior Officials | 1111-1161, 1171,1172, 1211, 1212, 1231, 1241-1243, 1251, 1254, 1255, 1258, 1259 | 1. Financial managers and directors (16%, n=253)<br>2. Managers and directors in retain and wholesale (11%, n=172)<br>3. Functional managers and directors n.e.c. (9%, n=142) |
| Other Professionals & Associate Professionals | 2111-2162, 2411-2455, 2471-2494, 3111-3133, 3411-3582 | 1. Programmers and software development professionals (5%, n=341)<br>2. Business and financial project management professionals (4%, n=248)<br>3. Management consultants and business analysts (4%, n=222) |
| Outdoor Trade Occupations | 5111-5119, 5311-5314, 5319, 5330, 8151-8159, 9111- 9129 | 1. Gardeners and landscape gardeners (22%, n=99)<br>2. Construction and building trades n.e.c. (21%, n=93)<br>3. Construction operatives n.e.c. (11%, n=50) |
| Sales & Customer Service Occupations | 7111-7220 | 1. Sales and retail assistants (38%, n=384)<br>2. Customer service occupations n.e.c. (11%, n=116)<br>3. Sales supervisors - retail and wholesale |

| | | |
|---|---|---|
| | | (9%, n=95) |
| Social Care & Community Protective Services | 1162, 1163, 1232, 2461-2469, 3221-3229, 3311-3319, 6134-6138, 6311-6312 | 1. Care workers and home carers (27%, n=285)<br>2. Welfare and housing associate professionals n.e.c. (11%, n=123)<br>3. Police officers (sergeant and below) (9%, n=91) |
| Teaching, Education & Childcare Occupations | 1233, 2311-2329, 3231, 3232, 6111-6117, 9232 | 1. Secondary education teaching professionals (14%, n=307)<br>2. Higher education teaching professionals (13%, n=285)<br>3. Teaching assistants (11%, n=274) |
| Transport & Mobile Machine Operatives | 8211-8239 | 1. Large goods vehicle drivers (20%, n=91)<br>2. Delivery drivers and couriers (19%, n=90)<br>3. Taxi and cab drivers and chauffeurs (14%, n=65) |

**Abbreviations:** n.e.c. = not elsewhere classified; * Limited to three most prevalent occupations per category to prevent declarative disclosure and due to large number of occupations across sample (*n*=412)

**Supplementary Figure 1.** Flow Diagram of Participant Selection

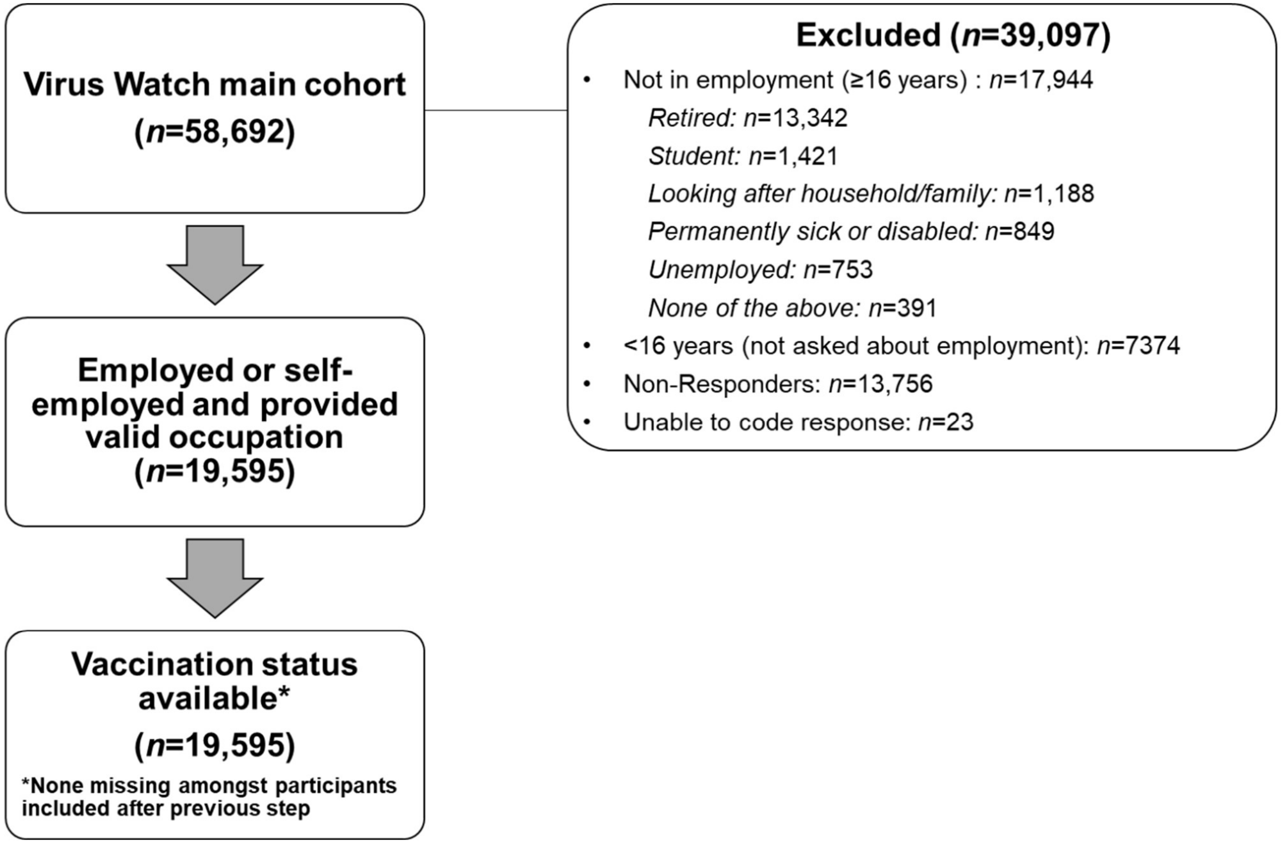

**Supplementary Figure 2.** Proportions and 95% Confidence Intervals for Clinical Vulnerability (a), Obesity (b), and Older Age (60+) (c) by Occupation

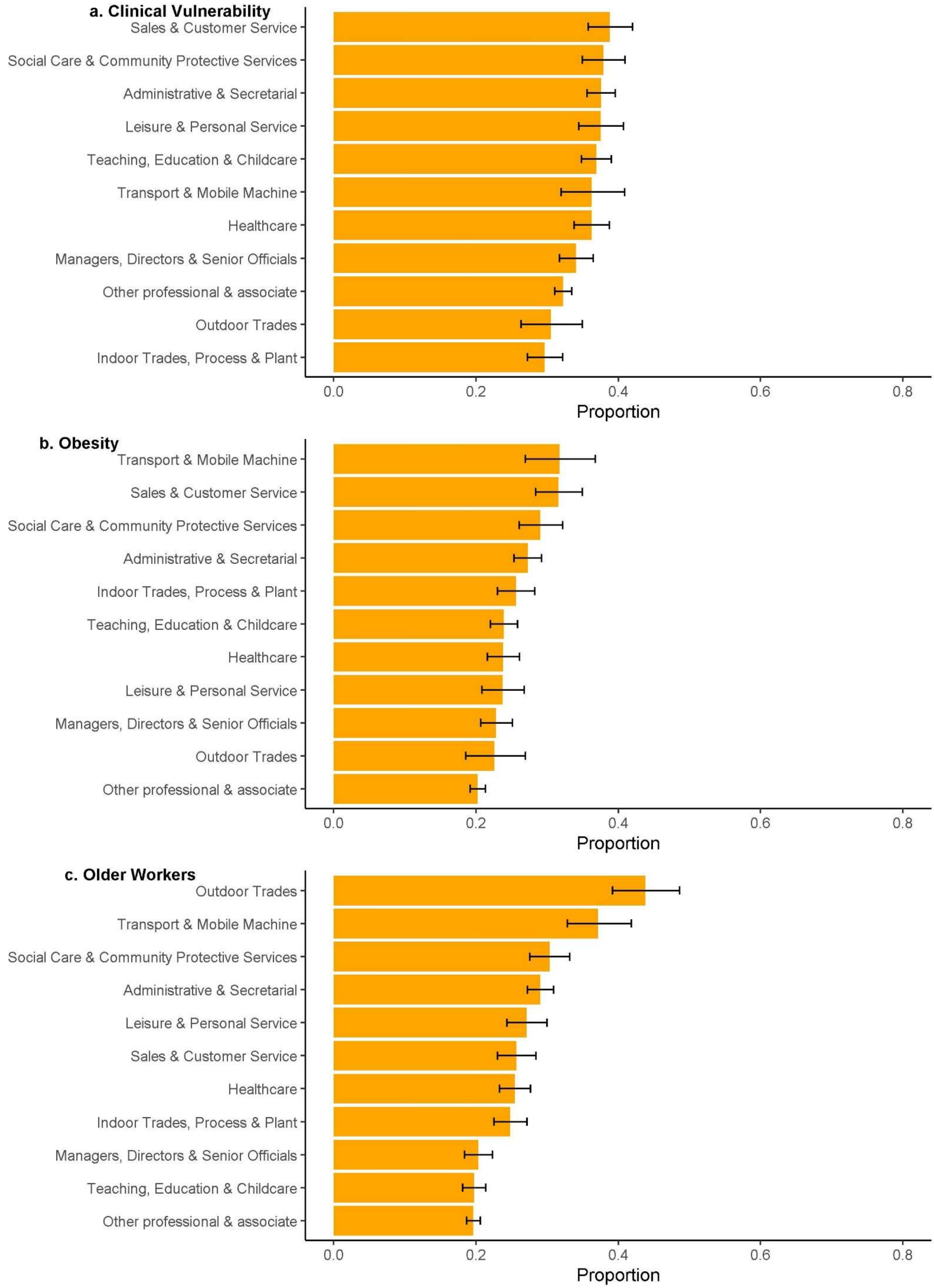

**Supplementary Figure 3.** Proportions and 95% Confidence Intervals for Vaccination Status by Occupation, Stratified by Obesity for First Dose (i), Second Dose (ii), and Third Dose (iii)

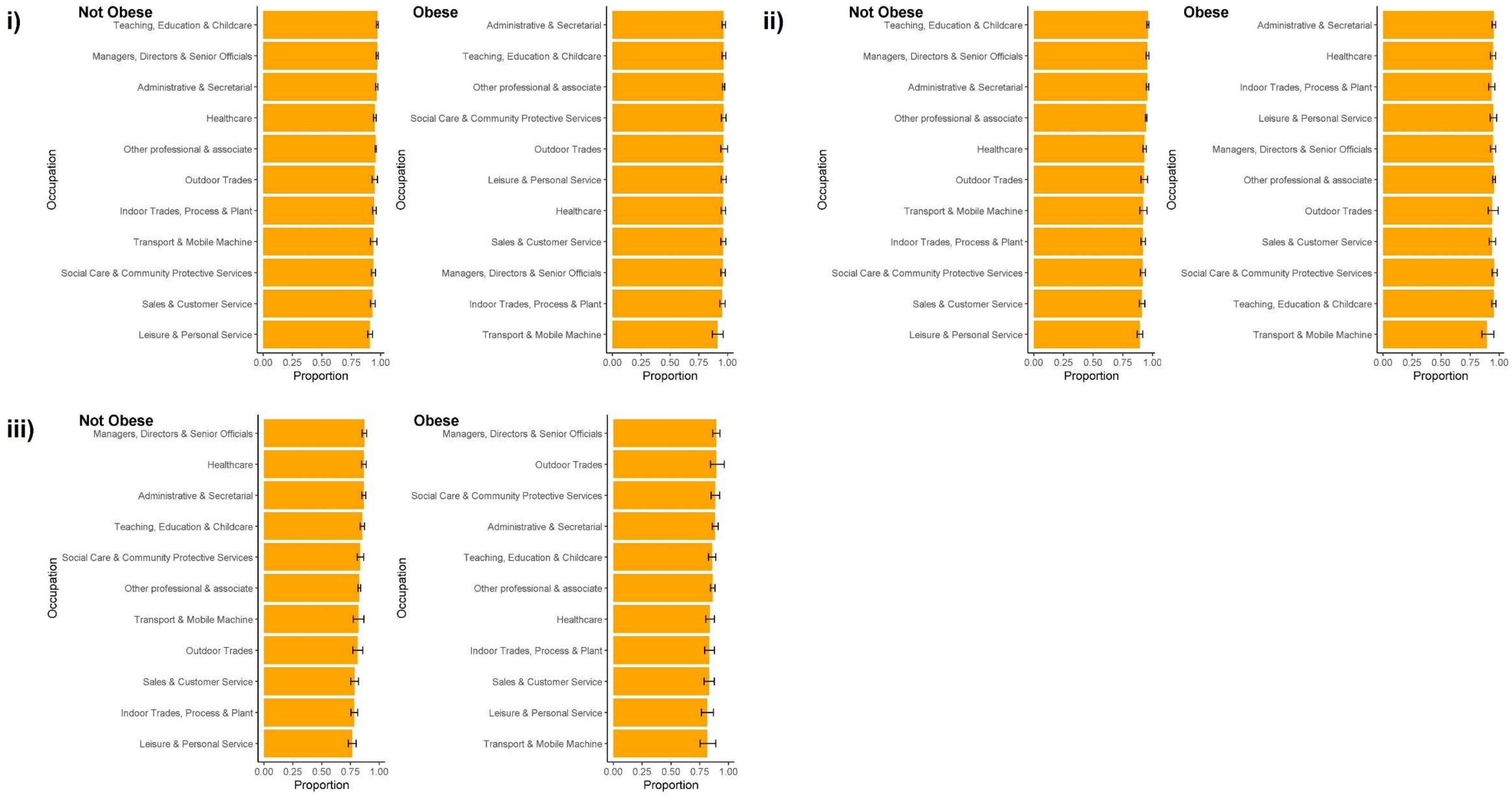
